# Development and Validation of the Participation Questionnaire for Preschoolers with Autism Spectrum Disorder: A Focus on Structural Validity, Internal Consistency, and Construct Validity

**DOI:** 10.1101/2024.02.13.24302559

**Authors:** Takuto Nakamura, Hirofumi Nagayama, Satoshi Sasada

**Affiliations:** Faculty of Health and Social Services, School of Rehabilitation, Kanagawa University of Human Services, Yokosuka, Japan

**Keywords:** Preschool Child, Autism Spectrum Disorder, Developmental Disabilities, Caregivers, Surveys and Questionnaires, Statistical Factor Analysis

## Abstract

**Aim:** To develop and validate the Participation Questionnaire for Preschoolers (PQP), a new disease-specific participation measurement tool for children with autism spectrum disorder (ASD)

**Methods:** In this cross-sectional study, questionnaires were distributed to caregivers of children through child development facilities and medical institutions. The sample comprised 180 children with ASD and 100 children with developmental disabilities without ASD. Additional participants with ASD were recruited through users of a developmental support website. Following item reduction based on the data from children with ASD, exploratory factor analysis was conducted, and Cronbach’s alpha coefficients were calculated. Correlations between PQP scores and age, symptom severity, sensory processing, and family functioning were evaluated to examine the construct validity. PQP scores for children with ASD were then compared with those for children with developmental disabilities.

**Results:** In total, 412 participants completed the questionnaire. Item reduction analysis led to the exclusion of seven items, revealing four factors. The PQP demonstrated high internal consistency and good construct validity, with observation of score differences based on the presence or absence of an ASD diagnosis.

**Conclusion:** The PQP appears to be an effective tool for children with ASD. However, further research is required to assess its longitudinal properties.

## Introduction

Autism Spectrum Disorder (ASD) is a neurodevelopmental disorder characterized by a wide range of features, including difficulties in forming interpersonal relationships, communication challenges, and repetitive patterns of behavior (American Psychiatric Association 2022). Generally, ASD becomes apparent in the early stages of development, suggesting that developmental experiences during early childhood have a significant impact in later life. Therefore, early childhood support is deemed crucial, and significant financial resources are allocated for this purpose to high-income countries (Lord et al., 2022).

Participation, as defined by the International Classification of Functioning, Disability and Health (ICF), refers to ‘involvement in life situations’ and is a widely accepted construct among professionals working with children with ASD (World health Organization, 2007). Within the ICF framework, participation is delineated into two dimensions: “capacity,” which indicates the extent to which an individual can participate under ideal conditions, and “performance,” which indicates the extent of participation in real-life situations. Furthermore, the ICF does not distinctly separate the constructs of “activity” and “participation.” While the domains of children’s participation, including self-care, interpersonal relationships, education, work, and leisure activities, are categorized into nine areas, the ICF does not explicitly differentiate which of these are strictly “activity” and which are “participation.” This ambiguity reflects the frequent overlap between activity and participation in real-life contexts.

Participation in early childhood and school-age is known to be restricted because of the unique disability characteristics associated with ASD (Askari et al., 2015). Research has shown that children with ASD encounter difficulties in various aspects of daily life participation, and these challenges have distinctive features compared with those faced by children with other disabilities (LaVesser & Berg, 2011; Schiariti et al., 2018). Occupational therapists, who are central professionals in supporting children with ASD, consider participation one of the most crucial outcomes (Boop et al., 2020). Therefore, they have been continuously developing and validating standardized tools to assess and utilize participation as an outcome measure in children with ASD.

To date, many participation measurement tools are generic and applicable to a wide range of conditions and are not confined to specific disabilities or diseases (Yee et al., 2017). These generic tools have enabled the investigation of how participation differs according to various disabilities and diseases, thus providing foundational knowledge for occupational therapists to support their participation (Askari et al., 2015). However, literature reviews examining tools for measuring participation in children with ASD emphasize the need for the development of tools tailored to the unique characteristics of ASD to adequately assess the impact of challenges, such as difficulties in adapting to new situations, social and communication difficulties, and repetitive interests and behaviors.

The earliest specific participation measurement tool for ASD, the Matrix for Assessment of Activities and Participation (MAAP), was developed for professionals to assess the activities and participation of children with ASD aged 3–6 years (Castro & Pinto, 2015). Although this tool offers the advantage of comprehensive participation measurement, it requires further research for item development and content validation. Golos et al. developed the Structured Preschool Participation Observation for Children with ASD [SPO-ASD], a tool used by professionals to assess participation in educational settings for children with ASD. This tool has been validated for content and reliability, providing a specific measure for participation in educational environments for children aged 4–6 years (Anat Golos et al., 2022). Additionally, the Picture My Participation-Chinese version (PMP-C) enables self-reported participation assessment for children aged 5–9 years through a photo-based interview with validated content (Li et al., 2023). While these tools offer numerous advantages, they also present challenges in terms of the time and expert involvement needed for implementation, which can be a barrier to their use in extensive research owing to human and time resource constraints (Lohr & Zebrack, 2009).

Nakamura et al. developed the Participation Questionnaire for Preschoolers (PQP), a questionnaire to be completed by caregivers (Nakamura et al., in press). The PQP is a participation measurement tool for children with ASD developed through interviews with caregivers and professionals, which measures participation in a disease-specific and comprehensive manner. It consists of 36 items that measure participation in a disease-specific and comprehensive manner. This easy-to-use, standardized measurement tool has the potential to allow for more efficient screening and monitoring of participation in clinical settings and to enable large-scale surveys to provide reliable evidence of participation in children with ASD. Thus, this study aimed to develop a PQP scale and investigate its structural validity, internal consistency, and construct validity.

## Methods

### Design

This cross-sectional study was conducted between December 2021 and February 2023. Data were collected through convenience sampling.

### Participants

Specifically, 56 child development centers and medical facilities across the country gave permission for collaboration with this study, and questionnaires were distributed by staff at the facilities to caregivers of children who used their services. Caregivers with children who met all of the following criteria were considered eligible: (1) preschool age between 36 and 83 months, (2) no diagnosis of a neurodevelopmental disorder other than the one being studied, and (3) a Developmental Quotient (DQ) of ≥50 as measured by the Wechsler Intelligence Scale for Children (WISC), the Wechsler Preschool and Primary Scale of Intelligence (WPPSI) (Wahlstrom et al., 2018), the Tanaka-Binet Intelligence Scale (TBIS) (Uno et al., 2014), or the Kyoto Scale of Psychological Development (KPSD) (Koyama et al., 2009), in accordance with Declaration of Helsinki. All caregivers received a document detailing the purpose, procedures, risks, and benefits of the study along with the mailed questionnaire. Caregivers indicated their consent by reading the document and returning the questionnaire by mail.

The children were categorized into two groups on the basis of the collected data: children with a diagnosis of ASD (ASD group) by a physician and children without a physician-established diagnosis of ASD (non-ASD group). The sample size for this study was based on the COnsensus-based Standards for the selection of health Measurement INstruments (COSMIN) criteria (Mokkink et al., 2018), and it was determined that a complete dataset with no missing PQPs for 180 children in the ASD group and 100 children in the non-ASD group was required, and data collection was continued with the goal of achieving this number. However, the possibility of a missing dataset for the ASD group arose, and a new data collection method was considered. This new approach utilized a mailing list maintained by a website for parents interested in child development and parenting, with approximately 300,000 registered users (Litalico Inc, 2023). Caregivers with children who met the aforementioned criteria and who had been diagnosed with ASD by a physician were invited to participate. Respondents were sent a questionnaire and informed consent was obtained using the aforementioned methods. Data were collected from child development offices and medical facilities from December 2021 to February 2023 and from respondents on the mailing list from January to February 2023. The study was approved by the ethical review board of the first author’s university (18-23-1).

### Measures (Instrumentation)

#### Participation questionnaire for preschoolers (PQP)

The PQP, a questionnaire administered to caregivers to measure the participation status of children with ASD, comprises 36 items. The response options are on a 5-point Likert scale; however, for this study, a “do not know” option was added to assess the understandability of the items. Higher scores denote a more favorable participation status of the children (Nakamura et al., 2023).

#### Social Communication Questionnaire Japanese Version (SCQ-J)

The questionnaire, designed to measure the severity of core symptoms of ASD, comprises 40 items and has demonstrated adequate reliability and validity (Uchiyama, 2015). It measures the severity of the core symptoms of ASD and has been shown to have sufficient reliability and validity. A preliminary validation of the cutoff values was conducted, and seven points were proposed as tentative cutoff values (Uchiyama, 2015). This is applicable to individuals with a mental age of ≥2 years. There are two types of test forms: “From Birth to Present” and “Present.” The former was utilized in this study. A higher score on this test indicates a greater severity of core symptoms.

#### Short Sensory Profile (SSP)

The SPP is a parent-response questionnaire comprising 38 items designed to assess children’s sensory processing abilities; it has high reliability and validity (Hirashima, 2016). Responses are given on a five-point scale ranging from 1 (never) to 5 (always). These domains include tactile sensitivity, taste/smell sensitivity, movement sensitivity, under-responsive/seeks sensation, auditory filtering, low energy/weak, and visual/auditory sensitivity. A higher score indicates more atypical sensory processing abilities.

#### Japanese Version of the Survey of Family Environment (SFE-J)

The SFE-J, designed to measure family functioning by assessing the environment both inside and outside the family, comprises 30 items and has adequate reliability and validity (Hohashi & Honda, 2012). Although there are several methods for calculating the SFE-J score, in this study, we calculated the overall satisfaction score, which was the average score for the items enquiring about the level of satisfaction with family functions. A higher score indicates better family function.

#### Demographic questionnaire

A demographic questionnaire was utilized to gather information about the respondents and their relationships with children diagnosed with ASD. The questionnaire covered various aspects, including the respondents’ educational background, family composition, annual household income, place of residence, and their children’s sex and age in months, comorbid diagnoses, and attendance at kindergartens and daycare centers. The IQ or DQ of the children was assessed based on the results of the WISC, WPPSI, TBIS, and KSPD; those with an IQ or DQ between 50–69 were classified into the intellectual Disability group, those between 70–84 into the borderline intelligence group, and those with an IQ or DQ of ≥85 into the standard intelligence group.

### Data analysis

The collected data were entered into a computer, and the responses to the questionnaires were cross-checked by the first author and two college students to minimize human error in data entry. Figure 1 shows the populations included in each statistical analysis. First, participants who were caregivers of children diagnosed with ASD were examined for ceiling and floor effects for each questionnaire item. Items with a ≥10% rate of the response “don’t know” and items with an item-total correlation coefficient of ≤0.3 were excluded. Next, an exploratory factor analysis was conducted for the responses from caregivers of children diagnosed with ASD who did not have missing PQPs. Items with communality of <0.3 and factor loadings of <0.3 were excluded from the scale, and Cronbach’s alpha coefficient was calculated to confirm internal consistency.

**Figure 1:**
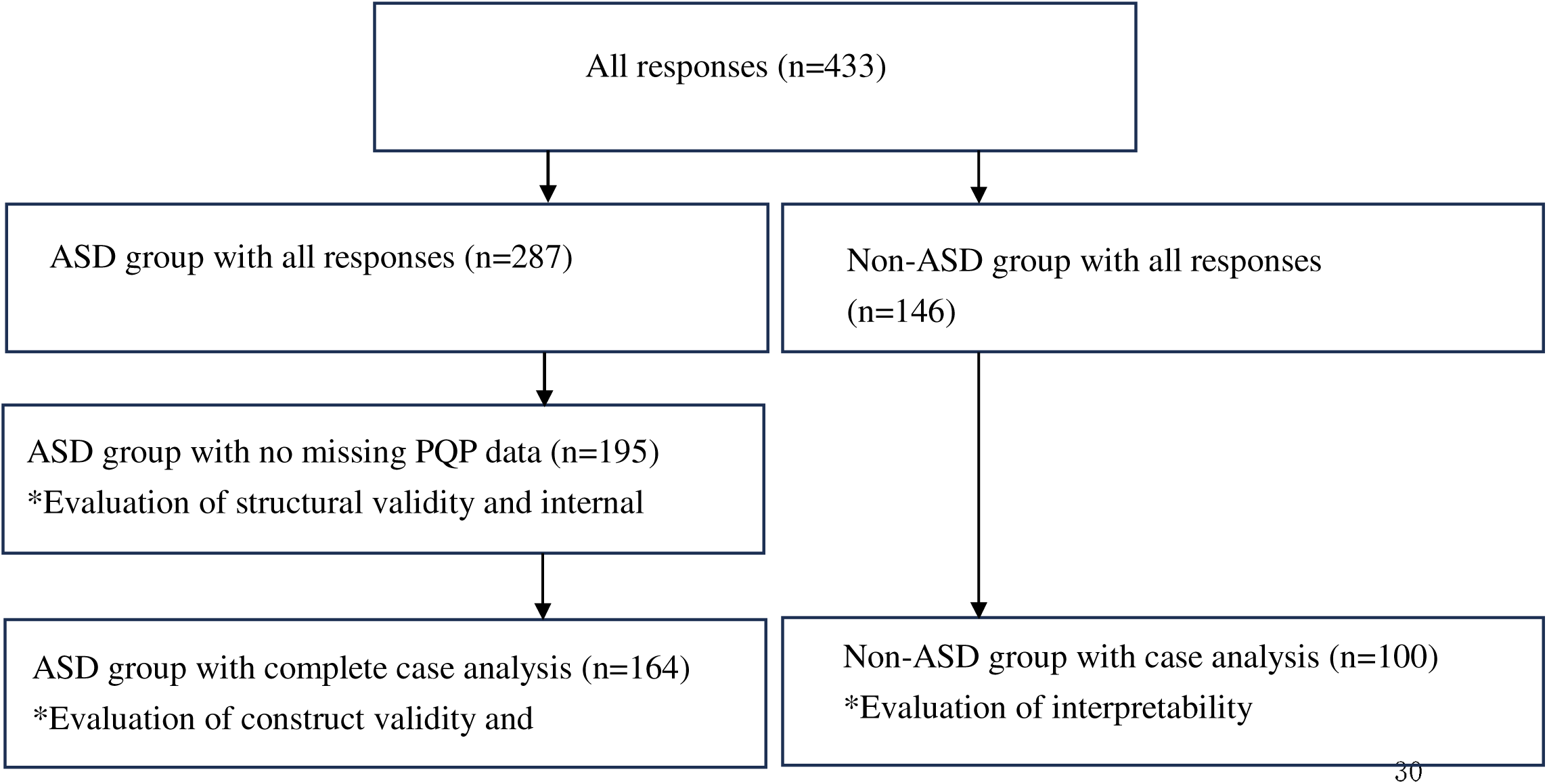
Group classification based on the presence of autism spectrum disorder and data completeness, with the statistical analyses ASD, autism spectrum disorder, PQP, Participation Questionnaire for Preschoolers

To test construct validity, Spearman’s rank correlation coefficients were calculated for responses from children with ASD without deficits in PQP, age, SCQ-J, SSP, and SFE-J, to test the following hypotheses:

1. Correlation between PQP score and age in months: Because no negative correlation between participation and age has been reported in early childhood (Gilboa & Fuchs, 2018; Khetani et al., 2015), we assumed a positive correlation between age in months and overall PQP scores, ranging from 0.0–0.4.
2. Correlation between the PQP and SCQ-J scores: Although the severity of core symptoms of ASD has been reported to be negatively correlated with out-of-school activities and friendships (Hilton et al., 2008; Orsmond et al., 2004), the relevance to the other domains of participation is not clear. Therefore, we assumed a negative correlation in the range of 0.2 to 0.7.
3. Correlation between PQP and SSP scores: As sensory processing is considered important in numerous participation domains, including education, play, leisure, social participation, eating, elimination, and sleep (Dunn et al., 2016; Ismael et al., 2018), we assumed a negative correlation in the range of 0.4 to 0.7.
4. Correlation between PQP and SFE-J: Because two studies on the relevance between participation of children with disabilities and family functioning suggest an association between participation and family functioning (Di Marino et al., 2018; King et al., 2006), we assumed a positive correlation in the range of 0.4 to 0.7.

Interpretability was tested by comparing data from the ASD group used in the factor analysis with data from the non-ASD group, which exhibited no PQP deficits. This approach was adopted under the premise that children with ASD and other neurodevelopmental disorders exhibit different patterns of participation, enabling clearer identification of ASD-specific patterns of participation and their needs compared to those for typically developing children. The PQP scores of the two groups were tested for significant differences using the Mann-Whitney U test. All statistical analyses were conducted using the IBM SPSS Statistics ver. 25.0 with a significance level set at 5%.

## Results

Valid responses were received from 433 participants. The response rate was 53.2%. Table 1 shows the demographic characteristics of the participants. The SCQ-J has previously been validated with a provisional cutoff point, with a recommendation to use seven points as the criterion (Uchiyama, 2015); however, this study was conducted with a small sample size and should be considered a preliminary criterion. In present study, some children in the ASD group did not meet this criterion; however, considering this background, we prioritized the diagnosis provided by a physician. Consequently we classified those with a diagnosis of ASD as the ASD group and those without a diagnosis as the non-ASD group. When the ASD and non-ASD groups with no missing data were compared, there were no prominent differences in many items. However, there were relatively large differences in the data collection method; region of residence; sex and education level of respondents; co-diagnosis of ADHD and neurodevelopmental disorders other than ID in children; and PQP, SCQ-J, SSP, and SFE-J scores. For other items, the differences were relatively small.

**Table 1.**
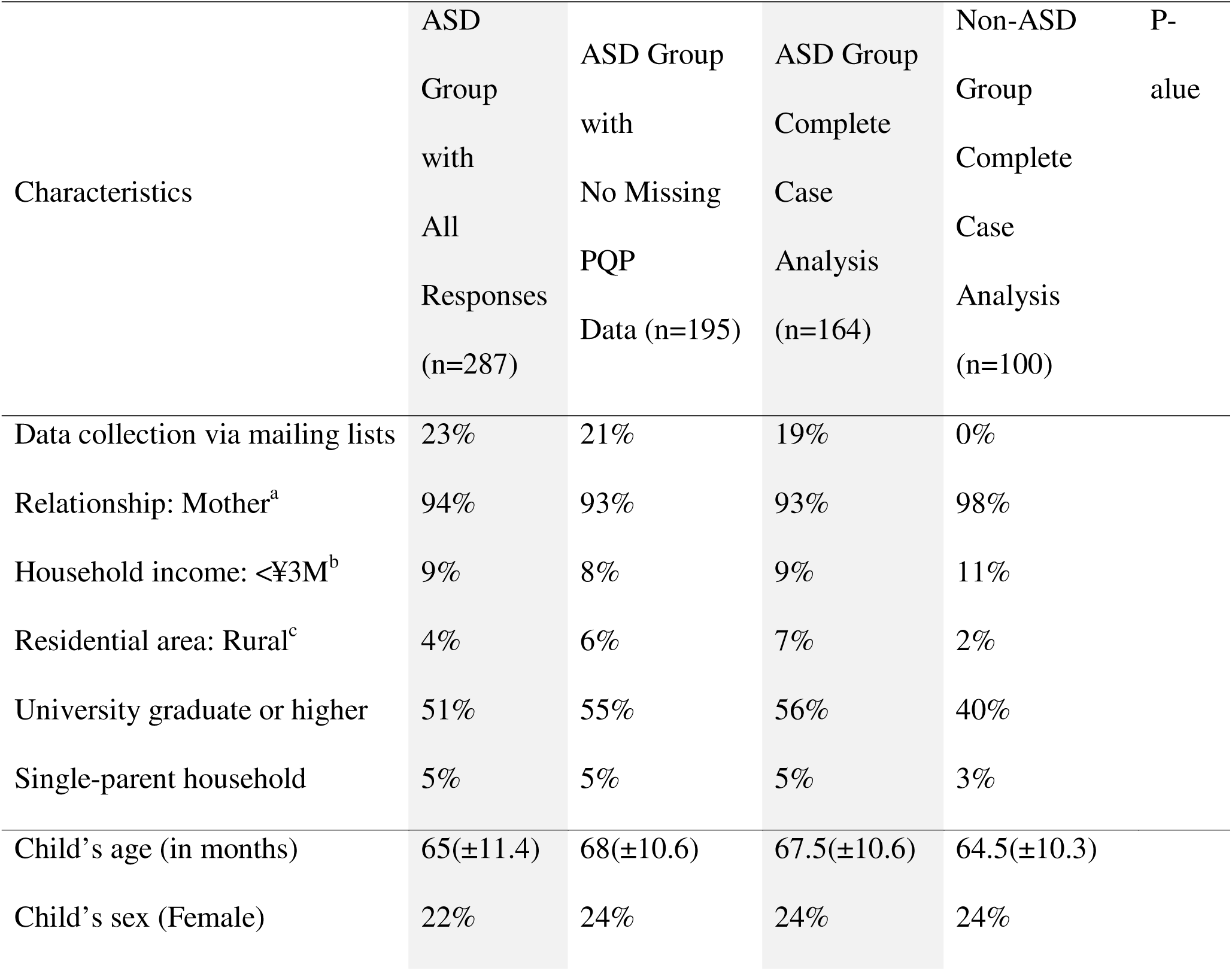
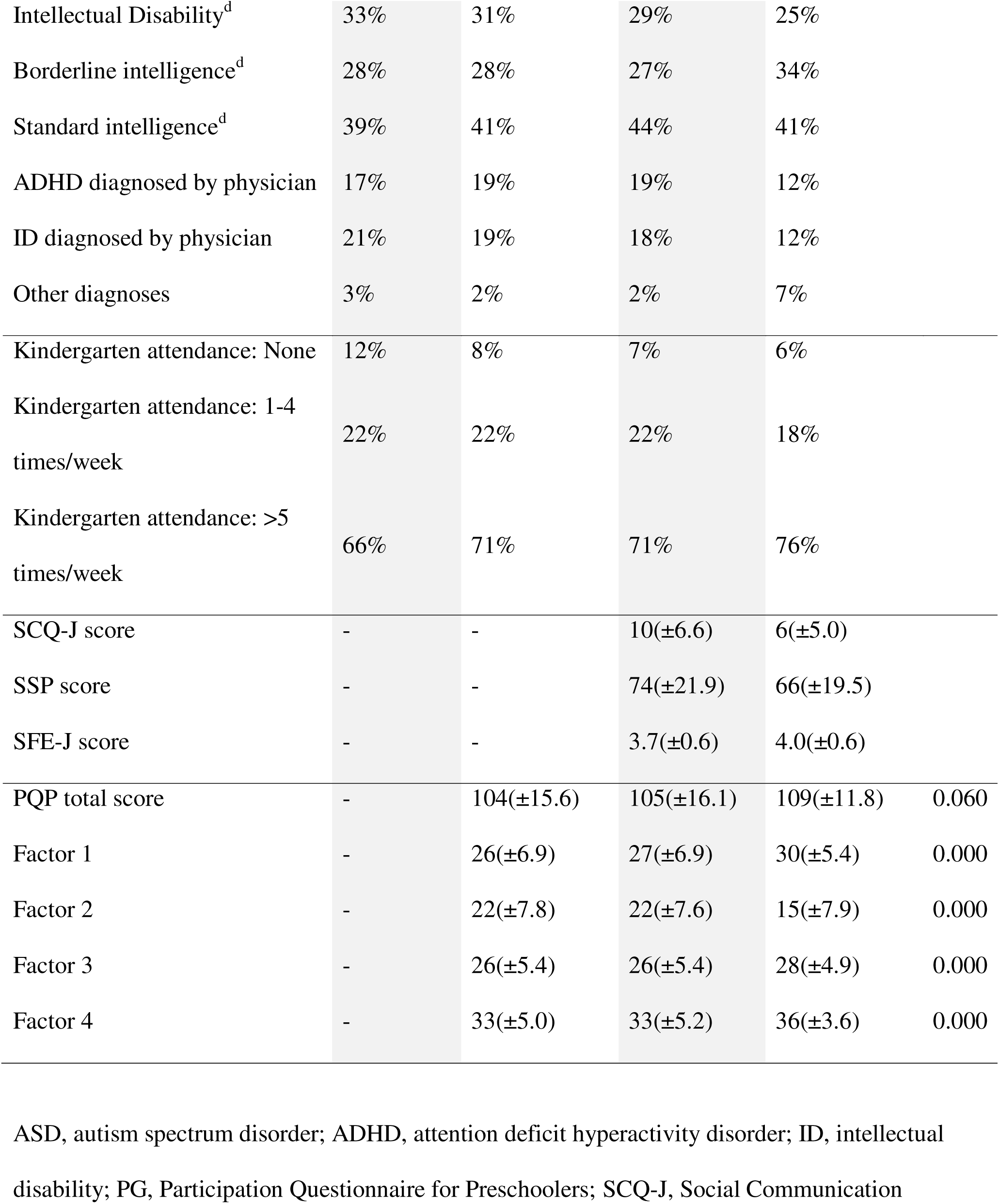
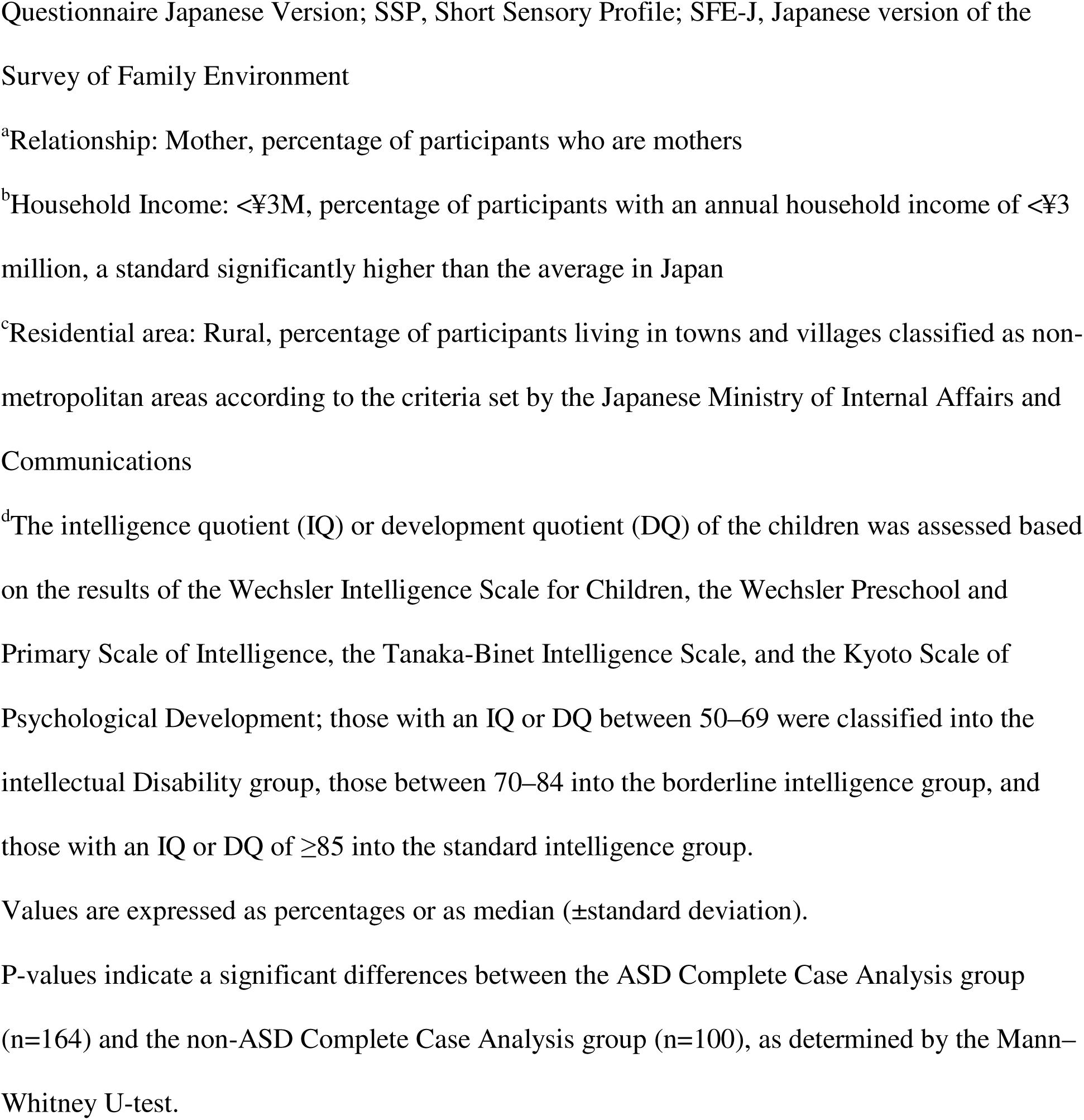
Demographic information for the caregivers and their children.

### Scale development and evaluation of structural validity and internal consistency

When the ceiling and floor effects were examined, one item exhibited a floor effect and 12 items displayed a ceiling effect. Next, for each of the 36 items, we calculated the percentage of respondents who chose “do not know” or provided no response, and we excluded item 11, i.e., “Has favorite lessons for learning,” because >10% of the total responses were as mentioned earlier. Furthermore, an item-total (IT) correlation analysis was conducted, and 12 items (1/3 of the total) had correlation coefficients < 0.3. Therefore, we adjusted the criterion for the correlation coefficient for the deletion of items to 0.2. Item 21 “Concerned about your child’s diet,” item 25 “Playing with your child can be exhausting,” and item 35 “Always seems to enjoy his/her time with the family” were deleted because they did not meet the criterion. Item 22 “Concerned about your child’s defecation” also showed a low correlation coefficient of 0.185; however, this item was retained because it was the only item related to elimination, a critical aspect that often requires support in the daily life of children with ASD based on the literature (O’Brien & Kuhaneck, 2019). The total number of items was 32.

Next, exploratory factor analysis was conducted using data from children with ASD without PQP deficiency. First, principal axis factoring was used, with the minimum eigenvalue set to 1, and the analysis was conducted using Promax rotation. The structure of the four factors was determined to be appropriate using a scree plot. Communality was adjusted to a criterion of 0.2 or less, aligning it with the IT correlation coefficient criterion, but no item fell below this criterion. Furthermore, when we reanalyzed the results using the Promax rotation of Principal Axis Factoring with the unique number of factors being 4, we found that item 20 “Opportunities to participate in community gatherings” (0.223) in factor 1, item 22” Concerned about my child’s defecation” (0.292) in factor 2, item 33 “Seems to enjoy mealtime” (0.256) in factor 3, and item 16 “Always has a teacher by his/her side to help them develop strengths” (0.245) in factor 4 had factor loadings of <0.3. However, item 22 was not deleted because of the aforementioned use.

Furthermore, we decided not to delete item 33 because it was the only item related to eating and because a large body of literature indicates that eating is a challenging activity for children with ASD (O’Brien & Kuhaneck, 2019). As a result, only items 16 and 20 were deleted, and exploratory factor analysis was conducted again; in addition to items 22 and 33, item 15 “Has a teacher that he/she likes” (0.281) in factor 1 was deleted because its factor loadings were below 0.3. After further analysis of the 29 items, the analysis was terminated because there were no other items with factor loadings of <0.3 besides items 22 and 33. The final factor patterns are presented in Table 2.

**Table 2.** Results of exploratory factor analysis.

| Item | 1 | 2 | 3 | 4 |
| --- | --- | --- | --- | --- |
| Plays with friends | 0.82 | -0.05 | 0.04 | -0.07 |
| Has close friends (Talks a lot, plays together a lot, etc.) | 0.81 | 0.02 | -0.12 | -0.11 |
| Enjoys class activities in cooperation with children of the same age | 0.80 | 0.03 | -0.01 | 0.05 |
| Learns social skills with friends | 0.78 | 0.09 | 0.05 | 0.00 |
| Always feels safe with friends | 0.65 | -0.08 | -0.11 | 0.14 |
| Participates in games with winners and losers (Relay, Sugoroku, etc.) | 0.63 | 0.02 | 0.19 | -0.04 |
| Learns how to use stationery (At home, kindergarten, etc.) | 0.40 | -0.04 | 0.28 | 0.09 |
| Difficulty in sending your child to preschool or kindergarten | -0.09 | 0.85 | 0.16 | -0.05 |
| Suffering from lack of specialized support for children with disabilities (Lack of facilities, reluctance to go to facilities, etc.) | 0.05 | 0.78 | 0.05 | -0.06 |
| Concerned about your child's sleep | -0.04 | 0.73 | 0.05 | 0.03 |
| Physically attacking a friend (Biting, hitting, etc.) | 0.13 | 0.66 | -0.10 | -0.00 |
| Concerned about changing your child's clothes | 0.03 | 0.55 | -0.17 | 0.14 |
| Reluctant to seek medical attention at a hospital | 0.00 | 0.40 | -0.04 | -0.02 |
| Concerned about your child's defecation | -0.06 | 0.30 | -0.07 | 0.07 |
| Helps around the home | 0.01 | -0.08 | 0.74 | -0.19 |
| Changes clothes by himself/herself | -0.06 | 0.06 | 0.65 | 0.15 |
| Finishes getting dressed | 0.11 | 0.04 | 0.64 | -0.03 |
| Seems to like helping out | 0.07 | -0.07 | 0.58 | 0.00 |
| Seems to like to keep up appearances<br>(Wash hair, brush teeth, etc.) | -0.08 | 0.04 | 0.45 | 0.13 |
| Engages in active physical play | 0.03 | -0.03 | 0.43 | 0.07 |
| Seems to enjoy mealtime. | -0.09 | -0.03 | 0.24 | 0.19 |
| You can take your child to a gathering of relatives | 0.02 | 0.08 | -0.12 | 0.72 |
| Plenty of places to go out with family | 0.08 | -0.05 | 0.05 | 0.63 |
| Time at home with them is always a pleasure | -0.13 | 0.06 | 0.06 | 0.56 |
| Can take your child on a trip that involves an overnight stay | 0.05 | 0.00 | 0.11 | 0.52 |
| Has someone other than his parents who is always there to help<br>him in his/her time of need | -0.05 | 0.05 | -0.08 | 0.48 |
| Neighbors outside of his or her immediate family are always nice<br>to him/her | 0.21 | 0.00 | -0.07 | 0.40 |
| Always seems to enjoy his/her time with the family | 0.05 | -0.14 | 0.20 | 0.34 |
| Can play alone for more than 15 minutes | -0.12 | -0.02 | 0.24 | 0.31 |

Factor 1 was interpreted as a factor related to social interactions with friends and learning in group life and was named “friendship and education.” Factor 2 was interpreted as a factor related to the distress that families face when living with children with ASD and was named “Family Satisfaction.” Factor 3 was interpreted as a factor related to the performance of daily activities, such as self-care and play, and was named “daily living and independence.” Factor 4 was interpreted as a factor related to leisure with family and participation in the community and was named “leisure and community life.” Together, all factors contributed 47.7% of the explained variance. The correlations between each factor and the total scores are presented in Table 3. To examine internal consistency, the alpha coefficients of each subscale were calculated, and the values of 0.883, 0.804, 0.752, and 0.738 were obtained for Factors 1, 2, 3, and 4, respectively. The alpha coefficient for all items was 0.826.

**Table 3.** Results of hypothesis testing for construct validity.

|  | **Total** | **Factor 1** | **Factor 2** | **Factor 3** | **Factor 4** |
| --- | --- | --- | --- | --- | --- |
| Child's age<br>(in months) | 0.215** | 0.237** | 0.122* | 0.113 | -0.014 |
| SCQ-J | -0.608** | -0.616** | -0.229* | -0.487** | -0.277** |
| SSP | -0.415** | -0.373** | 0.006 | -0.483** | -0.387** |
| SFE-J | 0.149** | -0.010 | -0.169 | 0.331** | 0.513** |

\*\*p < 0.01, \*p < 0.05; SCQ-J, Social Communication Questionnaire Japanese Version; SSP, Short Sensory Profile; SFE-J, Japanese version of the Survey of Family Environment

### Evaluation of construct validity

As shown in Table 3, three of the four hypotheses were supported.

1. Correlation between PQP score and age in months: A positive correlation within the hypothesized range was observed.
2. Correlation between PQP and SCQ-J scores: A negative correlation within the hypothesized range was observed.
3. Correlation between PQP and SSP scores: A negative correlation within the hypothesized range was observed.
4. Correlation between PQP and SFE-J: No positive correlation was found with the total PQP score, as hypothesized. However, there was a positive correlation with the scores for factor 3 and factor 4.

### Evaluation of interpretability

As shown in Table 1, ASD children scored significantly lower on factors 1, 3, and 4 and significantly higher on factor 2. Significant differences were found in all factors.

## Discussion

The present study aimed to develop a PQP scale and investigate its structural validity, internal consistency, and construct validity. Our findings reveal that the PQP had excellent structural validity, internal consistency, and construct validity. In the assessment of structural validity, four factors were identified, most of which showed no correlation. In the validation of internal consistency, each factor showed a good Cronbach’s alpha coefficient. This indicates that each PQP factor is capable of measuring distinct domains of participation. Given that three out of the four hypotheses tested for construct validity were supported, the PQP is generally expected to exhibit good construct validity. The correlation between PQP and SFE-J scores was lower than anticipated. This discrepancy might be attributed to the fact that previous studies (Di Marino et al., 2018; King et al., 2006) did not exclusively focus on ASD children, and that the present study used an external family functioning scale, which also concentrated on environmental factors surrounding the family, in contrast to the internal family functioning scale used in previous studies, which was centered on intra-family relationships (Nakamura et al., 2024). In addition, in the interpretability test, ASD children scored significantly lower for factors 1, 3, and 4, whereas children without ASD scored significantly lower for factor 2. While there are few studies comparing participation among young children with ASD and those with other neurodevelopmental disorders, the findings of this study align with those of previous studies (Askari et al., 2015) that indicate challenges in friendships and independence in ADLs for children with ASD. In addition, higher scores for factors related to family satisfaction were identified for children with ASD in the present study. This suggests that children with an ASD diagnosis are more likely to receive early professional support than are those with an undiagnosed condition. However, the present study did not explicitly measure the onset of professional support, and it is unclear how this unmeasured variable may have affected the results. Therefore, future studies should analyze the timing and quality of post-diagnosis support and its impact on family satisfaction in more detail.

Next, we discuss the characteristics of the PQP developed in this study by juxtaposing it with other ASD-specific participation measurement properties. First, as mentioned earlier, existing disease-specific participation measurement tools for ASD all assume professional involvement. In contrast, the PQP, to the best of our knowledge, is the first caregiver-completed participation measurement tool. Its independence from professional intervention not only reduces the time burden of intervention but also facilitates its application in large surveys. Second, the PQP provides a comprehensive measure of participation. Some previous participation measurement tools have focused on participation in specific settings, such as educational settings (Golos et al., 2022), or items have been developed without validating content relevance or comprehensiveness (Castro & Pinto, 2015). A convenient and comprehensive measure of young children’s participation may be useful for screening participation limitations in practice. In conclusion, the PQP was found to be a convenient, disease-specific participation measurement tool for children with ASD, demonstrating some degree of reliability and validity.

Finally, we discuss the limitations and future challenges. A limitation of this study is that the PQP scores may have decreased due to coronavirus disease 2019 lockdown measures and other factors (Children and Families Agency, 2023). Therefore, the standard scores and factor structure of the PQP should be reexamined in the future. In addition, the children in the SCQ-J study had cognitive development of at least 2 years of age, and it is possible that some children did not meet this criterion (Uchiyama, 2015). Furthermore, several items were deleted during the creation of the scale, necessitating future verification to determine whether the comprehensiveness of the items has been maintained. Longitudinal reliability and responsiveness validity, which are important measures of patient-reported outcomes, need to be verified for the PQP to be used to determine the validity of interventions in the future. By addressing these issues, the PQP may evolve into a useful tool for supporting children with ASD in early childhood. Furthermore, given the tendency of previous participation studies on young children to be conducted cross-sectionally in relatively small populations, fully leveraging the convenience of the PQP and measuring longitudinal changes in participation through large-scale surveys is an extremely important attempt to build evidence of participation among children with ASD.

## Conclusion

In this study, seven items were eliminated during the development of the PQP scale. The results showed that the PQP exhibited good measurement properties in terms of structural validity, internal consistency, and construct validity, indicating that the PQP may be a useful tool to comprehensively assess the participation of children with ASD; however, the comprehensiveness of the items and longitudinal validation of the measurement properties are future issues. Further studies should focus on the suitability of the questionnaire to other populations and its feasibility for intervention planning, by detecting children with a high burden of disease on everyday life.

## Data Availability

The participants of this study did not give written consent for their data to be shared publicly, so due to the sensitive nature of the research supporting data is not available.

